# GLP1R gene expression is associated with metabolic and mental health effects in a phenome-wide drug repurposing and safety analysis

**DOI:** 10.1101/2025.10.13.25337903

**Authors:** Jefferson L. Triozzi, Hua-Chang Chen, Fatih Mamak, Zhihong Yu, Otis D. Wilson, T. Alp Ikizler, Brian R. Ferolito, Kelly Cho, John Michael Gaziano, Ran Tao, Alexandre C. Pereira, Adriana M. Hung

## Abstract

Glucagon-like peptide-1 receptor agonists (GLP1RAs) are transformative therapies for diabetes and obesity, yet their long-term clinical effects are incompletely understood. To address this, we conducted a phenome-wide association study (PheWAS) to assess potential therapeutic and adverse effects of GLP1RAs using genetic drug proxies near the GLP1R locus. Fifteen variants associated with GLP1R expression in the Genotype-Tissue Expression (GTEx) project were tested against 1,204 phecodes in the Million Veteran Program (MVP, n = 464,626), with replication and meta-analysis within the UK Biobank (n = 449,349) and the Vanderbilt BioVU (n = 118,130). In the MVP, GLP1RA proxies showed expected metabolic benefits, including lower risk of type 2 diabetes (OR = 0.966, 95% CI [0.957-0.974], p = 4.55 × 10^−14^] and obesity (OR = 0.978, 95% CI [0.969-0.986], p = 6.94×10^−7^). Protective effects were also found for chronic venous insufficiency, obstructive sleep apnea, cancer of the esophagus, orthopnea, and diabetes complications (retinopathy, nephropathy, retinopathy). However, associations with mental health disorders were cohort-dependent (anxiety disorders in MVP OR = 1.02, 95% CI [1.012-1.0316], p = 1.07 × 10^−5^, BioVU OR = 0.960, 95% CI [0.938-0.981], p = 3.12 × 10^−4^). The results reinforce the therapeutic promise of GLP1RAs for metabolic diseases while underscoring the need for cautious monitoring of potential mental health effects.

## Introduction

Glucagon like peptide 1 receptor agonists (GLP1RAs) are widely used for the management of diabetes and obesity, with proven cardiovascular and mortality benefits [1,2]. Their therapeutic applications have expanded to include heart failure and diabetic kidney disease [3,4]. However, long-term clinical data are limited [5]. Current post-market surveillance systems face challenges such as underreporting, incomplete post-market studies, and limited ability to assess long-term or rare adverse effects in diverse real-world populations [6]. To address this gap, genetic variants influencing GLP1R expression, which naturally mimic the drug effects of GLP1RAs, can be used to investigate therapeutic and adverse outcomes [7].

Phenome-wide association studies (PheWAS) enable the identification of clinical diagnoses associated with the genetically proxied drug effects [8]. The genetically determined expression of the GLP1R gene is associated with metabolic traits, including adiposity and glycemic control, supporting its use as model for studying GLP1RA-related pathways [9]. By systematically by linking GLP1R genetic variants to the broad spectrum of clinical diagnoses in electronic healthcare records, PheWAS uncovers potential therapeutic benefits, safety signals, and other genotype-phenotype relationships that may not emerge during traditional clinical trials [10]. Because genetic variants are naturally occurring and allocated randomly at birth, they simulate a lifelong exposure to drug-like effects, exceeding the duration of traditional clinical trials [11]. Together, these features make PheWAS suited for identifying safety signals (i.e., SafeWAS). This framework extends traditional pharmacovigilance by proactively detecting adverse drug signals in large populations.

In this study, we examined the relationship between genetically determined expression of the GLP1R gene and a broad spectrum of clinical phenotypes, defined by phecodes in large biobank cohorts. This approach evaluated whether genetic proxies for GLP1RAs replicated the anticipated metabolic benefits, like reduced obesity and diabetes, while also uncovering additional genotype-phenotype associations that could signal potential therapeutic repurposing or safety concerns.

## Methods

### Study Design

The PheWAS used 15 genetic variants associated with GLP1R gene expression from the Genotype-Tissue Expression (GTEx) project as the exposure and up to 1,204 phecodes derived from the Million Veteran Program (MVP) biobank as the primary outcomes. Genetic variants were selected in an unbiased manner based on their association with GLP1R gene expression across multiple human tissue types, as described below. Clinical phenotypes were defined using phecodes mapped from International Classification of Diseases (ICD) codes available in the electronic health records (EHR) [12,13]. Statistically significant findings from the MVP were replicated in the UK Biobank (UKB) [14,15] and Vanderbilt University Biobank (BioVU) [16,17], based on de-identified genetic and phenotypic data. In all cohorts, participants provided informed consent for genetic and health record or survey use, and study protocols adhered to ethical guidelines for human subjects research. The current study is approved by the VA Central Institutional Review Board and the MVP Publication and Presentation Committee. The current study follows the STrengthening the REporting of Genetic Association Studies (STREGA): An Extension of the STROBE Statement [18].

### Study Population

The study population comprised participants from the MVP, a large biobank established by the U.S. Department of Veterans Affairs [19]. Participants provided blood samples for genotyping, completed health surveys, and consented to ongoing clinical data integration. Genotyping was done using a custom Affymetrix Axiom biobank array with quality control measures and imputation using the 1000 Genomes Project, as described elsewhere [20,21]. The replication and meta-PheWAS cohorts included the UKB and BioVU. UKB is a large prospective cohort of adults aged 40 to 69 from across the United Kingdom and this analysis used PheWAS data with TOPMed-imputed genotype data, as described elsewhere [14,22]. BioVU is a DNA biobank derived from clinical blood samples linked to a de-identified electronic medical records at Vanderbilt University Medical Center, as described elsewhere [12].

### Exposure

Based on a prior study, 15 genetic variants associated with GLP1R gene expression were identified by meta-analyzing their effects across 49 tissue types using RNA-sequencing data from the Genotype-Tissue Expression (GTEx) database, version 8 (**Supplemental Table 1**) [23]. In brief, GTEx provides eQTL data from 15,201 RNA-sequencing samples from 49 tissue types from 838 individuals [24]. Genetic variants within 1 Mb of the GLP1R transcription start site were evaluated, reflecting potential local regulatory effects. The effect of each genetic variant on GLP1R expression was meta-analyzed across all 49 tissue types using a random-effects model that weighed each tissue’s normalized transcript per million value (TPM) for GLP1R. This approach accounted for tissue-specific GLP1R expression, assigning greater weight to tissues with higher transcript abundance, such as the pancreas, reflecting their greater biological relevance to GLP1RA pathways. After selecting significant genetic variants and removing correlated genetic variants in linkage disequilibrium r^2^ < 0.2, this yielded 15 significant and independent GLP1R genetic variants associated with GLP1R expression.

### Outcomes

Clinical phenotypes from VA MVP were defined using phecodes, which map International Classification of Diseases (ICD) codes to disease traits [25]. Cases for each phenotype were defined by having at least two instances of the diagnosis codes on separate dates, while controls were defined as participants without that phecode or any related phecodes within the same disease category. Phecodes were generated by the MVP Data Core [26]. Clinical phenotypes were replicated in the UKB and BioVU using analogous phecode definitions [15]. Diagnoses with less than 200 cases were excluded to avoid biased effect estimates in the logistic regression.

### Covariates

Covariates included age, sex, and the first 5 principal components (PCs) of genetic ancestry to account for population stratification. Age was calculated from the recorded date of birth at enrollment. Sex was self-reported by study participants. The genetic ancestry of participants was determined using the Harmonized Ancestry and Race/Ethnicity (HARE) method [27], which integrates genetically inferred ancestry with self-reported race and ethnicity. Analyses were limited to European genetic ancestry to minimize confounding effects from ancestral heterogeneity between the exposures and outcomes [28]. The PCs of ancestry were calculated using FlashPCA2 [29].

### Statistical Analysis

The main PheWAS analysis was performed using an established methodology wherein logistic regression models adjusting for age, sex, and the first 5 principal components were used to assess the association between each GLP1R genetic variant and clinical phenotypes in the VA MVP [12]. An FDR-adjusted p-value to control for multiple testing was applied using the Benjamini-Hochberg method with a significance threshold of < 0.05. To facilitate interpretation of PheWAS results, we generated a heatmap to visualize the direction and magnitude of each SNP’s effect on GLP1R expression across all 49 tissue types in GTEx. The heatmap shows how individual genetic variants influence GLP1R expression in tissue-specific contexts (**Supplemental Figure 1**). Analyses were formed in R v.4.3.3 using the *PheWAS* and *meta* packages [30].

### Replication and Meta-PheWAS

Significant genotype-phenotype associations identified in the MVP were evaluated for replication in the UKB and BioVU. Each index genetic variant was regressed on the phecode outcome under a logistic regression framework. To declare a successful replication, the association had to demonstrate effect estimates in the same direction as observed in the MVP and exceed a significance threshold of p < 0.05. We combined the effect estimates from all three cohorts (MVP, UKB, and BioVU) using a fixed-effects meta-analysis to yield a pooled association. A fixed-effects model was chosen because these cohorts were of similar ancestry composition and assumption of a common underlying genetic effect.

## Results

In the main PheWAS analysis of up to 464,626 individuals in the VA MVP (**Table 1**), there were statistically significant phenotypes within the endocrine, cardiovascular, respiratory, neurological, and mental health domains (**Supplemental Table 2**). We aligned the direction of SNP effects on GLP1R gene expression with the pharmacodynamics of GLP1RAs (**Supplemental Table 1**). For SNPs associated with increased GLP1R expression, we retained the observed association with clinical outcomes. For SNPs associated with decreased GLP1R expression, we reversed the direction of the association.

**Table 1.** Baseline characteristic of the study cohorts.

|  | **MVP** | **UKB** | **BioVU** |
| --- | --- | --- | --- |
| **N** | 464626 | 449349 | 118130 |
| **Age, mean (SD)** | 64 (14) | 57 (8) | 58 (22) |
| **Sex:** |  |  |  |
| **Female, N (%)** | 34413 (7%) | 251139 (56%) | 67206 (57%) |
| **Male, N (%)** | 430213 (93%) | 198210 (44%) | 50924 (43%) |
| **Comorbidities:** |  |  |  |
| **Diabetes Mellitus, N (%)** | 116712 (25) | 23733 (5) | 17454 (15%) |
| **Hypertension, N (%)** | 266974 (57) | 174139 (39) | 38781 (33%) |
| **Characteristics:** |  |  |  |
| **BMI, m/kg<sup>2</sup>, mean (SD)** | 30 (6) | 27 (5) | 30 (42.5) |
| **A1C %, mean (SD)** | 6.2 (1.2) | 5.5 (0.7) | 6.4 (2.0) |
| **Systolic blood pressure, mm Hg, median (IQR)** | 129 (16) | 139 (20) | 124 (24) |
| **Diastolic blood pressure, mm Hg, median (IQR)** | 76 (10) | 82 (11) | 73 (16) |

MVP—Million Veteran Program; UKB—UK Biobank, BioVU—Vanderbilt University Biobank.

Among the main analysis within the VA MVP, the strongest associations were found for *rs10305420*, a missense variant and a known pharmacogenetic marker of GLP1RA drug response. The *rs10305420* effect allele was associated with increased GLP1R gene expression across most tissues including the pancreas, stomach, and intestine (**Supplemental Figure 1**). For each copy of the T allele, *rs10305420* was associated with lower odds of type 2 diabetes (OR = 0.966, 95% CI [0.957-0.974], p = 4.55 × 10-14] and obesity (OR = 0.978, 95% CI [0.969-0.986], p = 6.94×10^−7^). Analogues metabolic phecodes were also significant (e.g., abnormal glucose, type 1 diabetes, morbid obesity, overweight, obesity, and other hyperalimentation). Protective effects were also observed with reduced chronic venous insufficiency (p = 1.99 × 10^−5^), obstructive sleep apnea (p = 9.52 × 10^−5^), and orthopnea (p = 1.27 × 10^−4^). The intronic variant *rs60567936*, which decreased GLP1R expression (each copy of the T allele reducing normalized transcript levels by −0.125 TPM), was associated with increased esophageal cancer. After reversing the direction of the effect to align with the proxied effect of GLP1RAs, we inferred that increased GLP1R expression was associated with decreased esophageal cancer (OR = 0.795, 95% CI [0.715-0.884], p = 2.40 × 10^−5^). The intronic variant *rs7744341*, which increased GLP1R expression (each copy of the C allele increasing normalized transcript levels by 0.0851 TPM), was associated with increased major depressive disorder in the (OR = 0.963, 95% CI [0.945-0.982], p = 1.91×10^−4^) and anxiety disorder (OR = 1.021, 95% CI [1.010-1.031], p = 7.48 × 10^−5^) within the MVP.

Several phenotypes in the metabolic domain discovered in the MVP successfully replicated in at least one of the external cohorts. Type 2 diabetes successfully replicated (BioVU OR□=□0.971, p□=□0.019). Type 2 diabetes with ophthalmic manifestations (UKB OR□=□0.910, p□=□0.044, BioVU OR□=□0.897, p□=□0.007) and type 2 diabetes with renal manifestations (BioVU OR□=□0.937, p□=□0.009) also successfully replicated. Across the endocrine, cardiovascular, respiratory, neurological domains, the direction and approximate magnitude of effect of GLP1R expression on clinical outcomes remained consistent (protective) in all three cohorts, although formal significance was often driven by the large MVP sample size (**Figure 2**). In contrast, major depressive disorder (OR□=□1.022, p□=□1.07□x□10^−5^ in MVP) and anxiety disorders (OR□=□1.021, p□=□7.48□x□10^−5^ in MVP) did not replicate in the UKB or BioVU. Notably, BioVU showed a reversal of effect for anxiety (OR□=□0.960, p□=□3.12□×□10^−4^) and a borderline protective association for depression (OR□=□0.974, p□=□0.060). This heterogeneity across cohorts weakened their formal significance in the meta-analysis.

**Figure 1.**
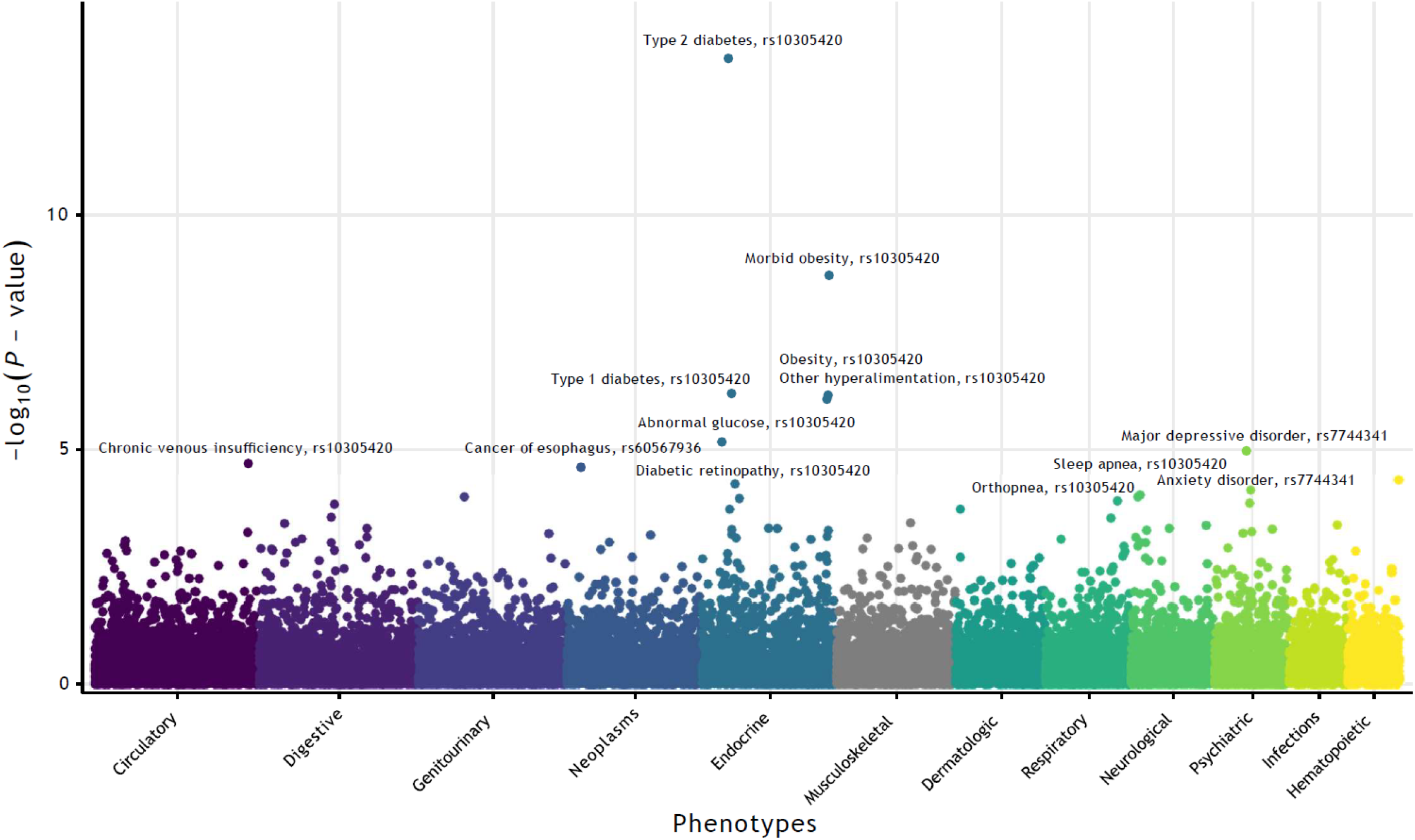
PheWAS Manhattan plot for the associations between 15 SNPs associated with GLP1R gene expression and 1,204 clinical phenotypes. Each point represents a SNP-phenotype association. Extended data are presented in **Supplemental Table 4**

**Figure 2.**
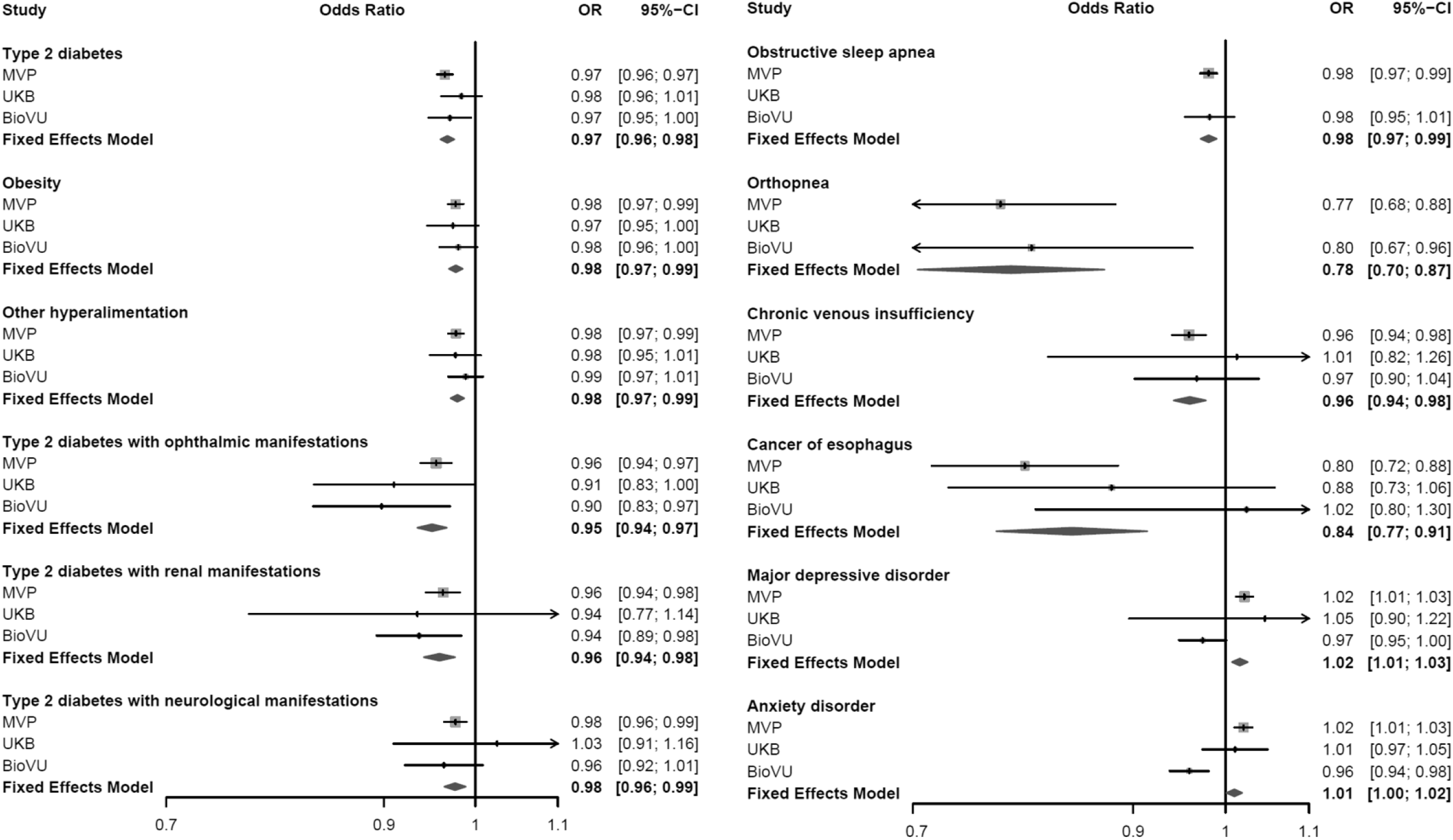
Forest plot for statistically significant genotype-phenotype associations in the Million Veterans Program that were replicated and meta-analyzed with the UK Biobank and Vanderbilt BioVU. MVP—Million Veteran Program; UKB—UK Biobank; OR—odds ratio; CI—confidence interval; DM—diabetes mellitus. Extended data are available in **Supplemental Table 5**. Analogous phecodes that were statistically significant but not pictured in this figure to reduce redundancy include: Morbid obesity 278.11, Abnormal glucose 250.4, Type 1 diabetes 250.1, Sleep apnea 327.3.

## Discussion

Our study reinforces metabolic benefits of GLP1RAs while highlighting potential mental health effects. The widespread use of GLP1RAs has brought attention to potential long-term adverse effects [5]. The associations between GLP1R gene expression and phenotypes provide a useful alternative to current post-market surveillance systems, by leveraging naturally occurring genetic variations to model drug effects and providing insights into drug efficacy and safety.

The genetic association between increased GLP1R expression and a reduction in odds of diabetes and obesity aligns with the well-known metabolic benefits of GLP1RAs, providing face validity to the genetic instruments. We also identified suggestive reductions in diabetes microvascular complications, which have been supported by prior observational data, clinical trials, and guideline consensus [31]. Notably, early concerns regarding GLP1RA use arose from the SUSTAIN 6 trial, which reported an increased incidence of diabetic retinopathy with semaglutide compared to placebo [32,33]. Subsequent pharmacovigilance studies and meta-analyses have produced conflicting findings, with many studies failing to confirm an increased risk of retinopathy. Similarly, we identified the effects of GLP1RAs extend to other conditions where weight loss is a cornerstone of treatment, such as obstructive sleep apnea and chronic venous insufficiency. GLP1RAs have shown promise in reducing the Apnea-Hypopnea Index and improving sleep quality in clinical trials [34]. Finally, the observed association with orthopnea represent a beneficial effect on congestive heart failure, consistent with known cardioprotective effects of GLP1RAs [35]. Several associations were driven by the genetic variant *rs10305420* which was associated with elevated endogenous GLP1R expression. However, as a pharmacogenetic variant, its effect allele is paradoxically associated with reduced weight loss and glycemic response to GLP1RAs such as liraglutide and exenatide [36,37]. This might suggest that naturally occurring variants like *rs10305420* may provide metabolic benefits via enhanced endogenous GLP1R signaling but exhibit diminished responsiveness to the therapeutic effects of GLP1RAs.

Our study also demonstrated a reduction in esophageal cancer risk, a finding of interest given prior conflicting evidence. Gastrointestinal side effects are the most commonly reported adverse effects of GLP1RA raising concerns for gastroesophageal complications [38,39]. Real-world evidence analyses have suggested that GLP1RA use indeed reduces the risk of obesity-associated cancers, including esophageal cancer [40]. However, other observational studies found that short-acting GLP1RAs were associated with increased risk of Barrett’s esophagus. These discrepancies may reflect differences in study designs, populations, residual confounding, and exposure duration.^22^ By contrast, genetic proxies represent sustained, lifelong GLP1R effects, mitigating many of these biases and providing data to support a reduction in long-term esophageal cancer risk.

Mental health conditions showed mixed results in our study, exemplified by opposing directions of effect in MVP and BioVU. Real-world data also point to a heterogeneous clinical landscape. A pharmacovigilance analysis of the FDA Adverse Event Reporting System identified a significant association between GLP1RAs and psychiatric adverse events, including nervousness, stress, eating disorders, and sleep disturbances [41]. Qualitative studies have descried diverse neuropsychiatric effects from GLP1RA, ranging from significant improvements in mood and mental health to reports of severe depressive symptoms and suicidal thoughts [42]. However, evidence from clinical trials is limited. Post hoc analysis of the Semaglutide Treatment Effect in People with obesity (STEP) trials found no clinically meaningful increase in the risk of depression or suicidal ideation with semaglutide compared to placebo in individuals with obesity, even suggesting a small reduction in depressive symptoms [1,43]. A real-world evidence analysis based on EHRs showed semaglutide was associated with a significantly lower risk of suicidal ideation compared to non-GLP1RA for obesity or diabetes [44]. Importantly, it remains possible that GLP1R signaling exerts context-dependent neuropsychiatric effects. Mental health outcomes may differ across cohorts, with age, sex, other medications, and underlying metabolic profiles, as well as other biological or environmental circumstances. The MVP cohort, composed of U.S. veterans, exhibits distinct demographic and clinical characteristics, including mostly men, a higher prevalence of comorbidities such as post-traumatic stress disorder, chronic pain, and other conditions [45].

## Strengths and Limitations

A strength of our study lies in the large-scale biobank analysis conducted within the MVP coupled with external replication and meta-analyses in independent cohorts from the UKB and BioVU. The selected GLP1R variants were identified through an unbiased approach without prior assumptions [23]. The random allocation of genetic variation at birth renders genetic instruments resistant to confounding and reverse causation biases. It also allows for the evaluation of a chronic drug effect over the course of a lifetime, longer than clinical trials or other observational studies [46]. For instance, a recent nationwide analysis in US veterans showed a decreased risk of psychotic and mood-related disorders in GLP1RA users. This study assessed benefits and risks of GLP1RAs across dozens of outcomes using a real-world pharmacoepidemiologic method that excels at detecting shorter-term drug-related signals but may remain vulnerable to residual confounding or prescribing practices [47]. This analysis However, translating genetic effects to clinical outcomes has inherent challenges. Genetic effects, while informative, could be smaller or larger in magnitude compared to the effects of pharmaceutical interventions. While patients experiencing adverse symptoms can discontinue treatment or adjust doses, genetic drug-like effects are lifelong and are unlikely to be modified. We also studied only genetic variants near the GLP1R gene, but there remains the potential for horizontal pleiotropy when a genetic variant exerts effects on phenotypes through pathways unrelated to GLP1R [48].

## Conclusions

In summary, genetic variants associated with GLP1R expression represent a lifelong, continuous exposure to a drug-like effect, providing insights into both the sustained benefits and potential risks of chronic GLP1RA. These genetic variants were associated with reduced risks diabetes and obesity, consistent with the established benefits of GLP1RAs, as well as suggestive benefits for other conditions such as obstructive sleep apnea, esophageal cancer, chronic venous insufficiency, diabetic ophthalmopathy, nephropathy, and neuropathy. The study further demonstrated a potential increased risk of depression or anxiety within the Million Veteran Program cohort, but disparate mental health effects in the Vanderbilt BioVU. Further research is required to clearly delineate which individuals are most likely to experience adverse mental health outcomes versus those who may benefit. Rigorous real-world pharmacovigilance to clarify the long-term clinical effects of GLP1Ras is needed.

## Acknowledgements

We are grateful to the MVP, UKB, and BioVU study participants for their contributions to science and all the enrolling sites listed in the Core Acknowledgement (**eAppendix**). J.L.T. had full access to all the data in the study and assume responsibility for data integrity.

## Disclaimer

This publication does not represent the views of the Department of Veteran Affairs or the United States Government. This research is based on data from the Million Veteran Program, Office of Research and Development, Veterans Health Administration.

## Data Availability

All genetic instruments are provided in the supplemental materials. The GTEx Portal is available from https://gtexportal.org/. The VA MVP individual level data will not be shared through dbGAP; the Office of Research and Development of the Department of Veterans Affairs needs to be contacted for access requests for that data.

