## Supplementary Materials for "GLP1R gene expression is associated with metabolic and mental health effects in a phenome-wide drug repurposing and safety analysis"

**Supplemental Content**

**Supplemental Figure 1.** Heatmap showing the effect size and direction of genetic variants associated with GLP1R expression across 49 tissue types.

**Supplemental Table 1.** Effect of genetic variants on GLP1R tissue when meta-analyzed across all 49 human tissue types in the GTEx, version 8.

**Supplemental Table 2.** All significant and nominal PheWAS findings in the Million Veteran Program.

**Supplemental Table 3 (Excel file attached).** Replication and meta-analysis of significant genotype-phenotype associations from the Million Veteran Program PheWAS within the UK Biobank and Vanderbilt BioVU.

**Supplemental Tables 4 (Excel file attached).** Complete main PheWAS results within the Million Veteran Program.

**Appendix.** VA Million Veteran Program: Core Acknowledgements for Publications.

**Supplemental Figure 1. Heatmap showing the effect size and direction of genetic variants associated with GLP1R expression across 49 tissue types.**

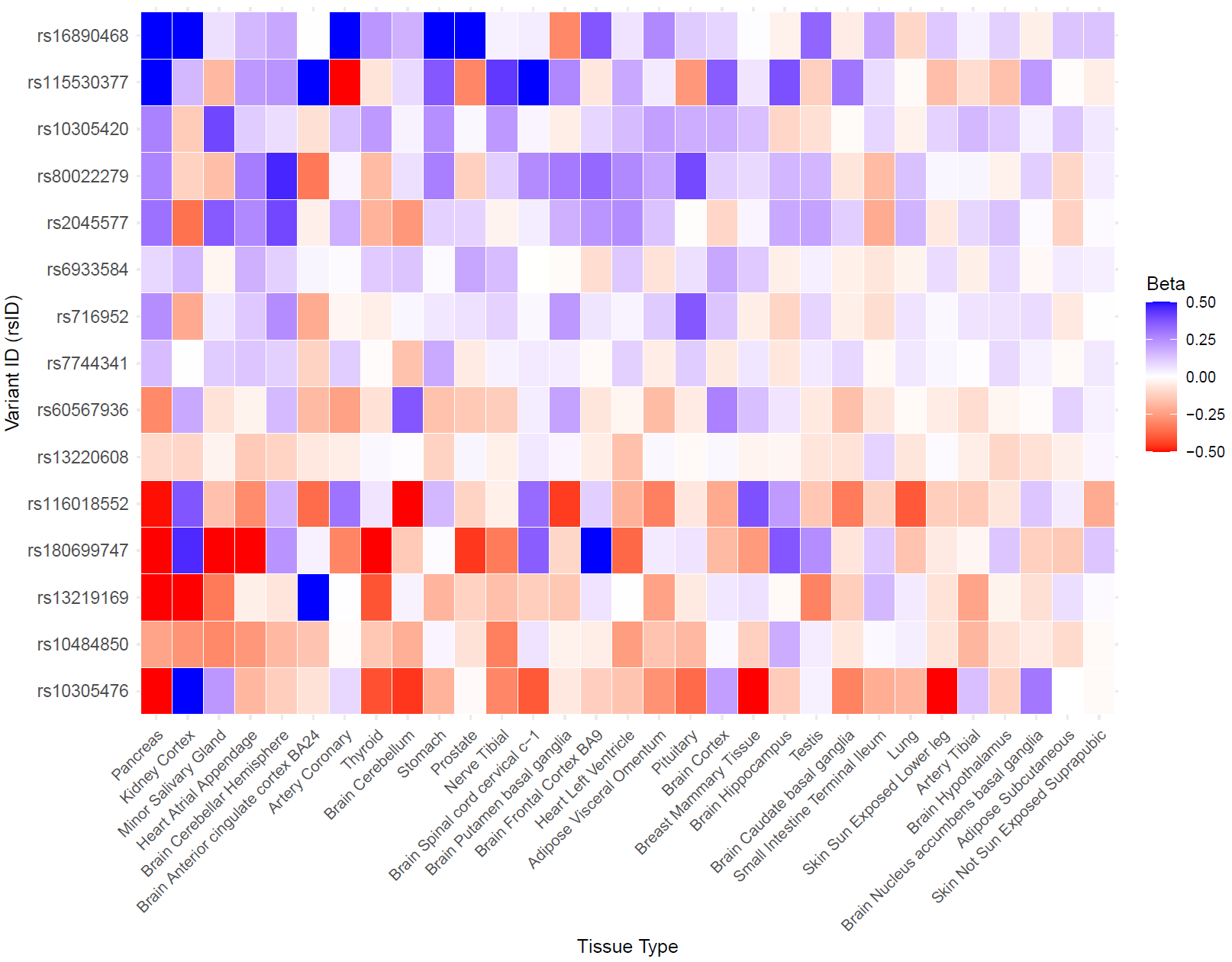

**Supplemental Table 1. Effect of genetic variants on GLP1R tissue when meta-analyzed across all 49 human tissue types in the GTEx, version 8.**

| **rsID** | **CHR** | **POS** | **REF** | **ALT** | **TSS Distance** | **Beta** | **SE** | **P-value** |
| --- | --- | --- | --- | --- | --- | --- | --- | --- |
| rs115530377 | 6 | 39264926 | G | A | 216128 | 0.284428282 | 0.090480884 | 0.001669302 |
| rs16890468 | 6 | 38237522 | C | T | -811276 | 0.226757225 | 0.07301479 | 0.001898715 |
| rs80022279 | 6 | 39397525 | T | C | 348727 | 0.172934222 | 0.055966245 | 0.002001742 |
| rs2045577 | 6 | 39615359 | T | C | 566561 | 0.165900278 | 0.049471281 | 0.000798061 |
| rs10305420 | 6 | 39048860 | C | T | 62 | 0.152523376 | 0.036476524 | 2.90E-05 |
| rs716952 | 6 | 39381589 | G | A | 332791 | 0.119934058 | 0.03202976 | 0.000180784 |
| rs7744341 | 6 | 39591175 | A | C | 542377 | 0.0850528 | 0.022002013 | 0.000110779 |
| rs6933584 | 6 | 38927373 | G | C | -121425 | 0.083491943 | 0.026901935 | 0.001912033 |
| rs13220608 | 6 | 39808404 | A | G | 759606 | -0.08120735 | 0.022217455 | 0.000257067 |
| rs60567936 | 6 | 39327729 | G | T | 278931 | -0.125151222 | 0.041185824 | 0.002376041 |
| rs10484850 | 6 | 39011665 | G | C | -37133 | -0.184525182 | 0.04068621 | 5.75E-06 |
| rs116018552 | 6 | 39128276 | G | C | 79478 | -0.252248207 | 0.077073094 | 0.001064713 |
| rs13219169 | 6 | 39051177 | G | A | 2379 | -0.28655591 | 0.065502766 | 1.22E-05 |
| rs10305476 | 6 | 39073305 | A | G | 24507 | -0.296309918 | 0.090057797 | 0.001001093 |
| rs180699747 | 6 | 38363354 | T | A | -685444 | -0.370303153 | 0.094071845 | 8.27E-05 |

CHR—chromosome; POS—position; REF—reference allele; ALT—alternative allele; TSS—distance of the genetic variant from the GLP1R transcription start site; SE—standard error.

**Supplemental Table 2. All significant PheWAS findings in the Million Veteran Program.**

| **Group** | **Description** | **Phecode** | **beta** | **SE** | **OR** | **p** | **FDR** | **n, controls** | **n, cases** | **Source** |
| --- | --- | --- | --- | --- | --- | --- | --- | --- | --- | --- |
| **Endocrine** | Type 2 diabetes | 250.2 | -0.0350 | 0.0046 | 0.966 | 4.55E-14 | 5.5E-11 | 289168 | 152865 | rs10305420 |
|  | Morbid obesity | 278.11 | -0.0433 | 0.0072 | 0.958 | 1.93E-09 | 1.2E-06 | 395388 | 46517 | rs10305420 |
|  | Type 2 diabetes with ophthalmic manifestations | 250.23 | -0.0453 | 0.0091 | 0.956 | 6.37E-07 | 2.0E-04 | 413690 | 27973 | rs10305420 |
|  | Obesity | 278.1 | -0.0227 | 0.0046 | 0.978 | 6.94E-07 | 2.0E-04 | 253541 | 173197 | rs10305420 |
|  | Overweight, obesity and other hyperalimentation | 278 | -0.0223 | 0.0045 | 0.978 | 8.44E-07 | 2.0E-04 | 228007 | 193176 | rs10305420 |
|  | Type 1 diabetes | 250.1 | -0.0519 | 0.0115 | 0.949 | 6.90E-06 | 1.4E-03 | 428091 | 16981 | rs10305420 |
|  | Abnormal glucose | 250.4 | -0.0249 | 0.0062 | 0.975 | 5.51E-05 | 8.3E-03 | 350420 | 67641 | rs10305420 |
|  | Diabetic retinopathy | 250.7 | -0.0344 | 0.0089 | 0.966 | 0.000113 | 1.2E-02 | 413490 | 29227 | rs10305420 |
|  | Type 2 diabetes with renal manifestations | 250.22 | -0.0373 | 0.0100 | 0.963 | 0.000191 | 1.8E-02 | 426060 | 22768 | rs10305420 |
|  | Type 2 diabetes with neurological manifestations | 250.24 | -0.0229 | 0.0066 | 0.977 | 0.00052 | 4.5E-02 | 387109 | 56634 | rs10305420 |
| **Mental Health** | Major depressive disorder | 296.22 | 0.0216 | 0.0049 | 1.022 | 1.07E-05 | 1.3E-02 | 297195 | 135516 | rs7744341 |
|  | Anxiety disorder | 300.1 | 0.0204 | 0.0052 | 1.021 | 7.48E-05 | 4.5E-02 | 306531 | 115776 | rs7744341 |
| **Neurologic** | Obstructive sleep apnea | 327.32 | -0.0194 | 0.0050 | 0.981 | 9.52E-05 | 1.2E-02 | 317120 | 121212 | rs10305420 |
|  | Sleep apnea | 327.3 | -0.0182 | 0.0047 | 0.982 | 0.000104 | 1.2E-02 | 290375 | 148430 | rs10305420 |
| **Respiratory** | Orthopnea | 513.32 | -0.2567 | 0.0670 | 0.774 | 0.000127 | 1.3E-02 | 457664 | 516 | rs10305420 |
| **Circulatory** | Chronic venous insufficiency | 456 | -0.0418 | 0.0098 | 0.959 | 1.99E-05 | 3.4E-03 | 419817 | 23962 | rs10305420 |
| **Neoplasms** | Cancer of esophagus | 150 | 0.2289 | 0.0542 | 1.257 | 2.40E-05 | 2.9E-02 | 457402 | 1933 | rs60567936 |

beta—effect size; SE—standard error; OR—odds ratio; FDR—false discovery rate-adjusted p-value.

**Appendix. VA Million Veteran Program: Core Acknowledgements for Publications.**

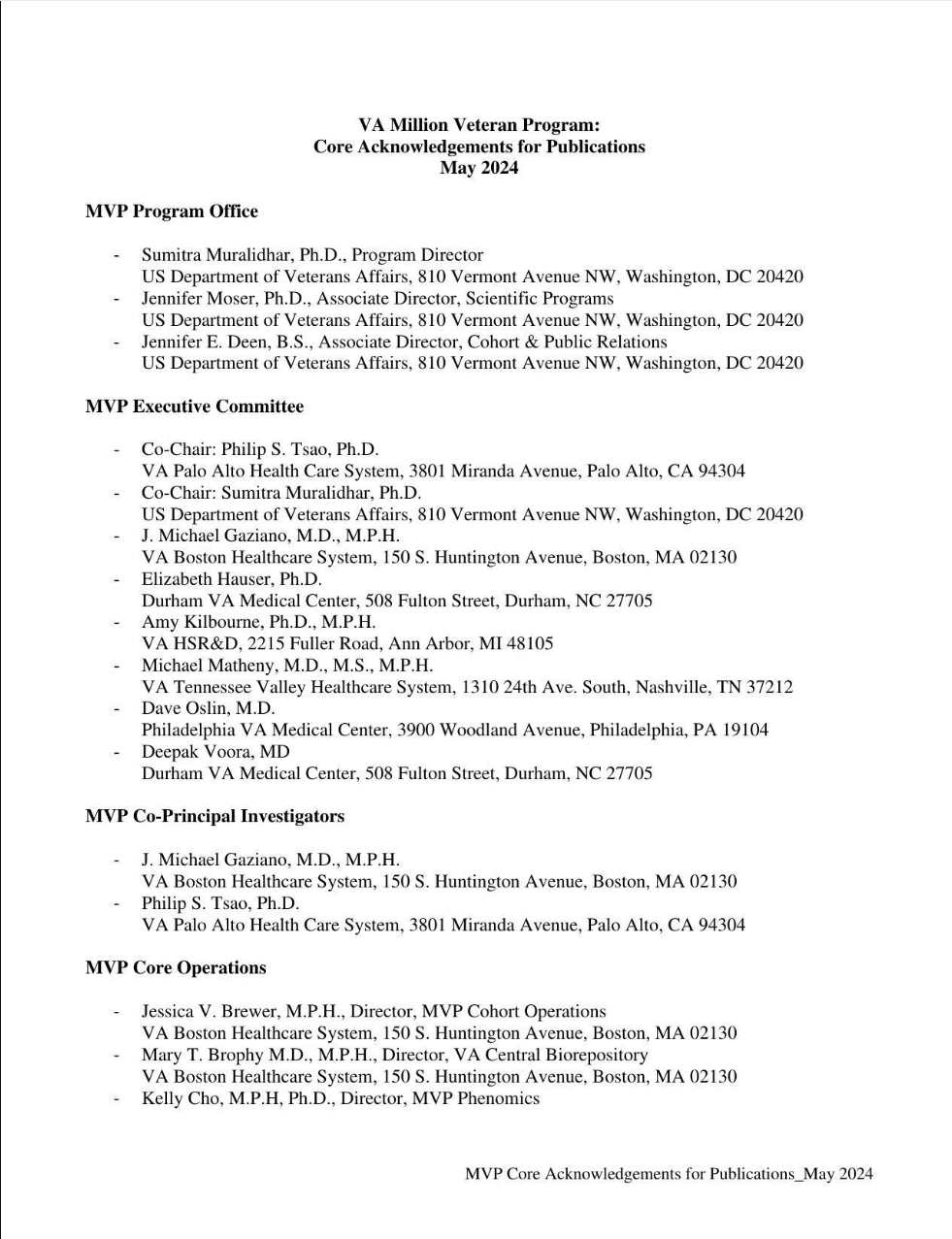

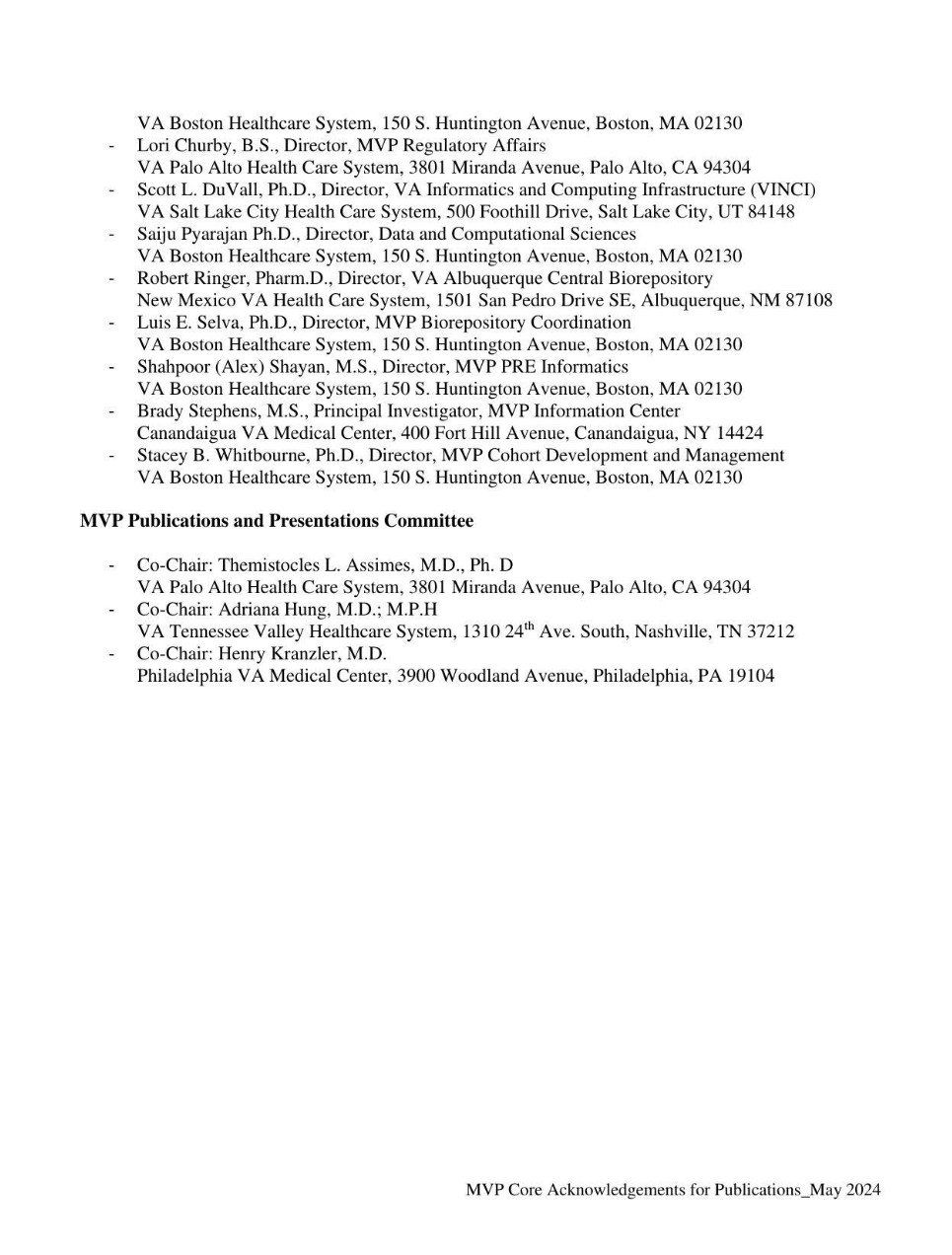
